# Quantification of the relative arm-use in patients with hemiparesis using inertial measurement units

**DOI:** 10.1101/2020.06.09.20121996

**Authors:** Ann David, StephenSukumaran ReethaJanetSureka, Sankaralingam Gayathri, Salai Jeyseelan Annamalai, Selvaraj Samuelkamleshkumar, Anju Kuruvilla, Henry Prakash Magimairaj, SKM Varadhan, Sivakumar Balasubramanian

**Author notes:** **Corresponding Author:** Dr. Sivakumar Balasubramanian Professor, Dept. of Bioengineering Christian Medical College Bagayam, Vellore Tamil Nadu, India – 632002.

## Abstract

**Background:** The most popular method for measuring upper limb activity is based on accelerometery. However, this method is prone to overestimation and is agnostic to the functional utility of a movement. In this study, we used an inertial measurement unit(IMU)-based gross movement score to quantify arm-use in hemiparetic patients at home.

**Objectives:** (i) Validate the gross movement score detected by wrist-worn IMUs against functional movements identified by human assessors. (ii) Test the feasibility of using wrist-worn IMUs to measure arm-use in patients’ natural settings.

**Methods:** To validate the gross movement score two independent assessors analyzed and annotated the video recordings of 5 hemiparetic patients and 10 healthy controls performing a set of activities while wearing IMUs. The second study tracked arm-use of 5 hemiparetic patients and 5 healthy controls using two wrist-worn IMUs for 7 days and 3 days, respectively. The IMU data obtained from this study was used to develop quantitative measures (total and relative arm-use (RAU)) and a visualization method for arm-use.

**Results:** The gross movement score detects functional movement with 50–60% accuracy in hemiparetic patients, and is robust to non-functional movements. Healthy controls showed a slight bias towards the dominant arm (RAU: 40.52º). Patients’ RAU varied between 15–47º depending upon their impairment level and pre-stroke hand dominance.

**Conclusions:** The gross movement score performs moderately well in detecting functional movements while rejecting non-functional movements. The patients’ total arm-use is less than healthy controls, and their relative arm-use is skewed towards the less-impaired arm.

**Clinical trial registry number:** CTRI/2018/09/015648

## Introduction

Impairment reduction following rehabilitation of hemiparetic patients often do not translate to an equivalent increase in functional arm-use^1–3^. This is because patients do not use their impaired limb to its full ability in their natural setting^4,5^. Quantitative measurement of arm-use in the natural setting is crucial for various reasons:- documentation, monitoring change over time, assessing compliance, motivating patients through feedback, etc. Traditional methods for arm-use measurement use structured questionnaires (e.g., Motor Activity Log (MAL)^6^) or observe spontaneous arm-use during selected activities in a clinical setting (e.g., Actual Amount of Use Test- AAUT^7^). These current approaches have several limitations: (i) questionnaires have reporter bias, and high variability in self-judgment of movements^8^; (ii) these are administered in a clinic for a short duration, thus do not necessarily measure arm-use during daily-living (e.g., bilateral arm use test^9^); and (iii) AAUT can only be administered once and cannot track arm-use over time. Therefore currently, there is lack of ecologically valid method to quantify arm-use in patients’ homes.

Wearable sensors help in the continuous, uninterrupted, objective measurement of arm-use in a natural setting^10–13^. Accelerometry-based activity counts that detect any upper-limb movement is currently the most popular measurement of arm-use^14–18^. However, they fail to isolate functional movements (e.g., ADL) from non-functional ones (e.g., detection of arm swing during ambulation), and attach equal importance to both types of arm movements. Quantifying arm-use by without distinguishing between functional versus non-functional arm movements while assumptions on the mobility status of patients can lead to overestimation of arm-use^19–21^. Some data-driven approaches using supervised or unsupervised learning algorithms to classify functional or non-functional movements through accelerometry yield high classification accuracy, but are restricted to specific tasks that were used in the laboratory setting^22–24^. A simple, elegant and general algorithm to detect functional arm-use of an upper-limb was proposed by Leuenberger et al. using a single IMU on the forearm^25^. This algorithm returns a binary signal – *gross movement score* – corresponding to the presence/absence of functional arm movements, which is insensitive to non-functional arm movements.

The current study explores the feasibility of using two wrist-worn IMUs for tracking arm-use at home in hemiparetic patients through a week-long pilot study. We employed the gross movement score for detecting functional movements to measure the relative upper-limb use in daily life. The duration of seven days was chosen based on the work of Trost et al., which recommended 3 to 5 days for monitoring activity using accelerometers^26^. Currently, there is no information on the types of functional movements detected by the gross movement score. Thus, we also validated the gross movement score against functional movements detected by a human assessor from video recordings of activities performed by healthy controls and hemiparetic patients wearing the IMUs.

## Methods

The study aimed to develop and validate a measure to monitor the relative arm-use in hemiparetic patients at home. The institutional review board of Christian Medical College Vellore (CMC) approved this study. The study has two parts: (a) an in-clinic validation of the gross movement score with a video recording of the patient’s arm use (IRB Min. No. 12321 dated 30.10.2019), and (b) a seven-day arm-use monitoring study at home using IMUs. (CTRI/2018/09/015648, IRB Min No. 11303 dated 18.04.2018).

### IMU watch

We designed an “IMU-watch” – a wrist-worn device housing a SEN – 14001 board (Sparkfun Inc.) containing a SAMD21 microprocessor with an inbuilt real-time clock, 9-DOF IMU (MPU9250, InvenSense-TDK Co.), MicroSD card slot, and a battery charging circuit. The IMU data and the real-time clock’s timestamp were logged at 50 Hz to an 8GB microSD card. Each subject wore two IMU-watches – one on each arm – whose real-time clocks synchronized to GMT +5.5 hours. An 800mAh rechargeable Li-Po battery powered the device for 12–15 hours on a full charge. The enclosures for the watches were 3D printed and were marked R and L for the right and left IMU-watches, respectively.

#### Gross Movement Score (*γ*)

Fig. 1 shows the processing pipeline for extracting the gross movement score time series *γ*[*n*] at time instant *n* from the raw IMU data. The Madgwick algorithm^27^ was used to estimate the yaw (*α*) and elevation (*β*) angles of the forearm with respect to an earth reference frame. These angles were used to compute *γ*[*n*] using a 2s long moving windows with 75% overlap (sampling time of *γ*[*n*] is 500ms).

**Fig 1:**
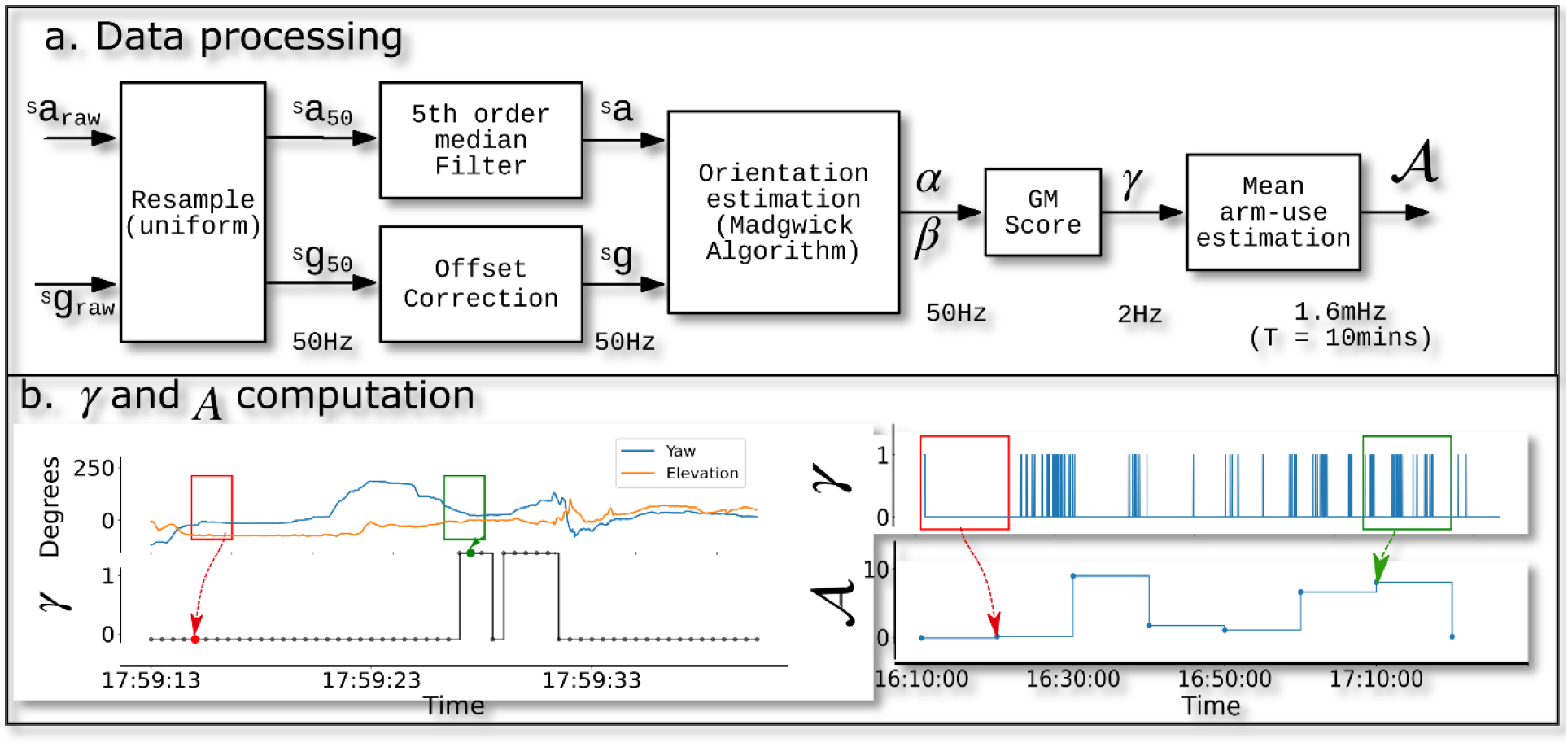
Data processing pipeline, and demonstration of the computation of *γ* and ***𝒜*** from a raw IMU data. a) The yaw (*α*) and elevation (*β*) angles are computed from raw acceleration(**a_raw_**) and gyroscope (**g_raw_**)at 50 Hz. Changes in *α* and *β* within the functional range is summarised as the gross movement score (*γ*) at 2Hz and mean arm-use (***𝒜***) at 1.6mHz (every 10 mins). b) In the left panel, changes in *α* and *β* in the red box is less than 30º so, *γ* = 0 (red dot b) but for the green box, total change in *α* and *β* is greater than 30º and *α* is within the functional range, hence *γ* = 1, (green dot). In the left panel, ***𝒜*** is computed as the percentage of total *γ* in 10 min window. Red and green boxes of width 10 min in the *γ* plots are used to compute ***𝒜*** at red and green dots respectively.

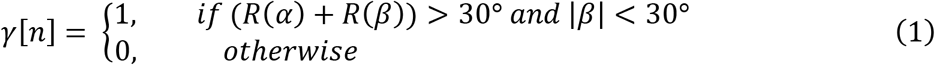

where the function *R*(·) computes the range of the input argument in a 2s window. The *γ*[*n*] assumes that functional movements occur within an elevation of ±30°; this is referred to as the *functional range* from here on.

#### Mean Arm-use (***𝒜***)

\Mean arm-use time series is computed as the percentage of time *γ*[*n*] = 1 in a non-overlapping 10min window (sampling time of ***𝒜***[*k*] is 10min; *k* is the time index for ***𝒜***). For example, ***𝒜*** of 40% at 9:00 AM implies that that the gross movement score was 1 for 4mins between 8:50 – 9:00 AM. For each subject, ***𝒜*** was computed for the left and right arms.

A scatterplot between the mean arm-use time-series of the two arms over an observation period (e.g., 12 hours of data from a day) is used to visualize relative arm-use. Here, the x-axis (***𝒜_x_[k]***) and y-axis (***𝒜_y_[k]***) correspond to the less- and more-impaired arm, respectively, for patients, and the dominant and non-dominant arm, respectively, for healthy controls. The points (0, 0) (i.e., zero mean activity for both arms) were excluded from visualization or any analysis carried out on the scatterplot. The spread of the points in this scatterplot allows us to quickly gauge the nature of arm-use. The distance of a point (*𝒜_x_*[*k*], ***𝒜_y_[*k*]***) from the origin 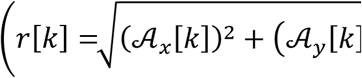 indicates the intensity of arm-use, which is a measure of how active the two arm were in given time window. The angle subtended by this point *θ*[*k*] = tan^−1^(^*𝒜_y_*^/_*𝒜_x_[k]*_) is a measure of the proportion of the use of the two arms in this time window. To visualize the intensity of arm-use *r*[*k*] as function of the relative proportion of use between the arms *θ*[*k*], a 1-dimensional curve *ρ*(*ϕ*) was generated from an arm-use scatterplot, which was computed as,

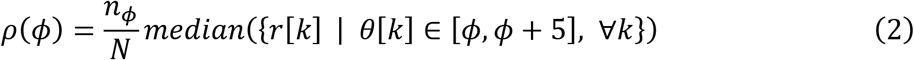

where, *N* is the total number of points in the scatterplot, *n*_*ϕ*_ is the number of the scatter points in the sector [*ϕ*, *ϕ* + 5], and *ϕ* ∈ {0, 5, 10, 15, ☦ 85}.

The area under the curve *ρ*(*ϕ*) is defined as the *total arm-use*, which was computed using the trapezoidal rule. The overall preference between the two arms called the *relative arm-use* (RAU) was computed as the following,

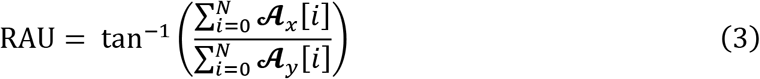

#### Validation of the Gross Movement Score

The validation of *γ* was carried out on healthy controls and hemiparetic subjects using the same data collection protocol, but with slightly different analysis procedures. The inclusion criteria for the patients with hemiparesis were: they should have (i) no severe cognitive deficits (Mini-Mental State Examination score (MMSE) higher than 25); (ii) Manual Muscle Test (MMT) grade higher than 2; (iii) age between 25 – 70 years; (iv) at least 30° elevation of the arm against gravity in the shoulder joint with the elbow extended; (v) 20° wrist extension against gravity; (vi) 10° finger extension (proximal metacarpophalangeal and interphalangeal) of at least one finger against gravity; (vii) ability to open the hand in any position to accommodate a small ball (diameter of

1.8cm) in the palm; and (viii) willingness to give informed consent. Patients who had pain while moving the upper-limb, and allergy to the plastic material used for the IMU-watch casing and straps were excluded from the study. Patients were recruited through the inpatient Occupational Therapy unit of CMC Vellore.

The inclusion criteria for healthy controls were: (i) no prior history of upper-limb movement problems due to neurological conditions; (ii) no current difficulty in upper-limb movements; (iii) age between 25 and 70 years; and (iv) willingness to give informed consent.

The participants were instructed to carry out a set of 15 tasks (see supplementary materials) while wearing two IMU-watches. These movements were recorded using a camera connected to a PC time-synchronized with the IMU-watches.

The videos from healthy controls were annotated independently by two biomedical engineers (not part of study) using custom-made software. The human assessors were instructed to identify gross movements within the functional range, ignoring the movements’ functional utility. This score marked by the human assessors is referred to as the *γ*^𝑣𝑖𝑑𝑒𝑜^[*n*] – a binary time series indicating the presence of a gross movement identified from the video at time *n*.

The videos from the patients were annotated independently by two Occupational Therapists (GS, RJS) using the same software. This analysis produced the functional activity (FA) score – a binary time-series indicating the presence of a functional movement at each instant.

The video annotations were repeated after ten days by the same assessors to estimate intra-assessor variability of the *γ^video^* and FA scores. The *γ^video^* and FA scores from the videos were down sampled to 2Hz to compare them individually with *γ* from the IMU-watches, through:

i. **Accuracy** defined as the percentage of data points in agreement, i.e., both *γ* and *γ*^video^ (or FA) are 0 or 1.
ii. **Gwet’s AC1** agreement statistic to account for chance agreement between two scores^28^.

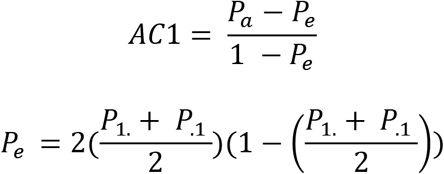

Where, *P_a_* is the sum of true positive and true negatives rates. *P*_1_. and *P*_.1_ is the marginal probability of *γ* and *γ^video^* (or FA) being true, respectively.

#### In-home arm-use assessment of hemiparetic patients

The inclusion/exclusion criteria for the healthy control and patients were the same as the one in the validation study described earlier. Additionally, patient’s residing outside 30km radius from CMC Vellore were excluded from study. The patients were recruited through the stroke clinic of CMC Vellore. Patients made a one-time visit to the Occupational Therapy unit for initial assessments. After obtaining informed consent, a modified AAUT^29^ was performed, followed by the upper-limb Fugl-Meyer Assessment, and the MAL. Each subject was given a pair of time-synchronized IMU-watches and a charger to take home, and was given a demonstration of donning the watches. Participants were directed to wear the watch marked R and L on the right and left wrist, respectively. They were instructed to wear the watches throughout their waking hours except when there was a risk of the watches coming in contact with water. Patients and healthy controls the watches for 7 and 3 days, respectively. At the end of this period, the watches were collected back for analysis.

## Results

### Validation of Gross Movement Score

Five patients with mild-to-moderate hemiparesis undergoing therapy at CMC Vellore, and 10 right-handed healthy controls participated in the study. The average age of hemiparetic patients was 38.4 years, while that of healthy controls was 23.3 years. Healthy controls performed all fifteen tasks, while patients completed the first 10 tasks (see supplementary material).

Fig. 2 shows four tasks and their corresponding *γ* scores (blue trace) and the movement identified by the human assessors (orange trace). The time instant corresponding to the video frames are shown at the bottom-right corner of the frame; these time instants are marked by a vertical red band in the adjacent graphs. The following observations can be made from this figure:

**Fig 2:**
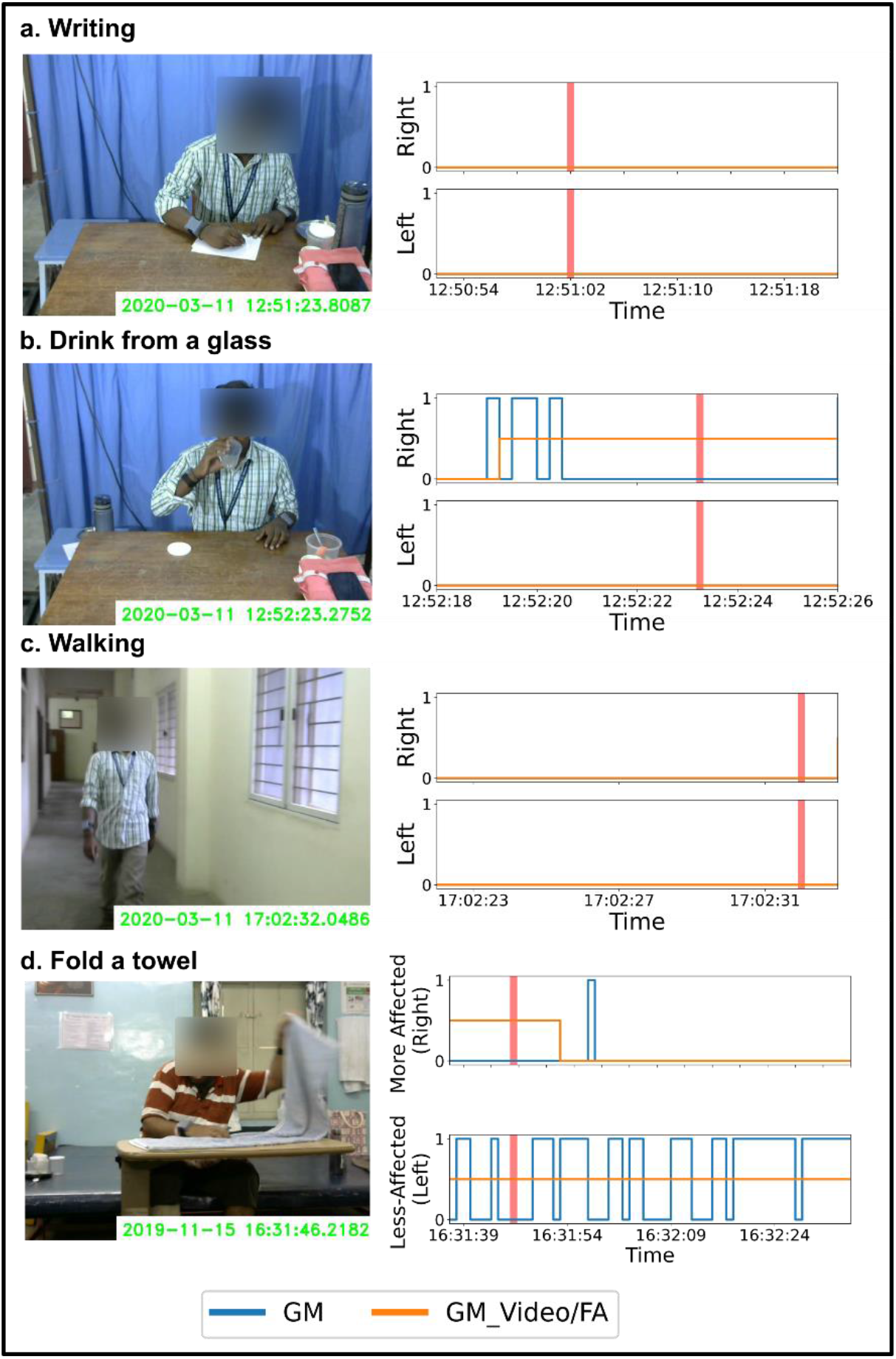
Video frames with FA/GM_Video (orange) and GM (blue) score during task execution. The red vertical lines correspond to the time at which the frame was captured for (a) writing, (b) drinking, (c) walking, (d) folding a towel. The *γ* identifies functional gross movements while accurately removing non-functional movements.

i. ***γ* does not identify functional tasks involving fine finger movements**, e.g. writing (Fig. 2(a)), typing on a keyboard, etc. This is seen as a 0 in both *γ^video^* and *γ*.
ii. **Some functional tasks are identified in fragments** as these tasks require the arm to move out of the functional range (±30°) used for computing *γ*. For example, when drinking from a glass (Fig. 2(b)), *γ* is marked as 0 at the time instant shown in the video frame as the arm is out of the functional range.
iii. *γ* **does not detect non-functional movements** like arm swing during ambulation (Fig. 2(c)); *γ* is uniformly zero during this activity.

Table 1 provides the summary of the comparison of *γ* with *γ^video^* and FA scores for the healthy control and patients, respectively. For patients, the accuracy of the IMU-watch in identifying functionally relevant tasks is between 53–58%, with a moderate AC1 agreement of 0.43–0.53 between *γ* and the functional activities identified by therapists. There was good consistency between and within the two therapists, with an inter- and intra-assessor AC1 agreement of 0.840 (0.072) and 0.878 (0.056), respectively.

**Table 1:** Agreement analysis (Accuracy, and Gwet’s AC1 scores) of *γ* with *γ^video^* /FA score of each arm for all subjects.

| Patients |  |  |  |  |  |
| --- | --- | --- | --- | --- | --- |
| Assessor | Assessment No. | Impaired Arm |  | Non-Impaired Arm |  |
|  |  | Accuracy | AC1 | Accuracy | AC1 |
| A(GT) | 1 | 56.766 | 0.525 | 54.298 | 0.454 |
|  | 2 | 56.806 | 0.523 | 54.078 | 0.450 |
| B(RJS) | 1 | 55.078 | 0.503 | 57.114 | 0.494 |
|  | 2 | 54.184 | 0.493 | 53.298 | 0.436 |
| Healthy |  |  |  |  |  |
| Assessor | Assessment No. | Left Arm |  | Right Arm |  |
|  |  | Accuracy | AC1 | Accuracy | AC1 |
| C | 1 | 81.5 | 0.80 | 70.7 | 0.67 |
|  | 2 | 79.6 | 0.78 | 68.3 | 0.65 |
| D | 1 | 65.5 | 0.62 | 45.2 | 0.35 |
|  | 2 | 71.3 | 0.69 | 57.2 | 0.50 |

For control subjects, the accuracy of *γ* was between 45–82%, and the AC1 agreement was between 0.35–0.80. This large variability in the control data was due to a relatively low inter-assessor AC1 agreement of 0.66975(0.07), even though the intra-rater agreement was good 0.82(0.053). It was observed that *γ* had better agreement with assessor C than assessor D, because assessor C had performed relatively fine-grained frame-by-frame marking than assessor D who used a coarse approach to annotate the video.

### In-Home Arm-Use Assessment

Five patients with hemiparesis due to stroke and five control subjects participated in this study. The patient demographics are given in Table 2. All patients were at least 3 months post-stroke, mild-to-moderately impaired as measured by the Fugl-Meyer Assessment (FMA), with MAL scores that appeared to be related to their impairment level. For the AAUT, patients could complete all tasks in the no-choice condition, but all patients except P1 showed some level of arm non-use during the spontaneous condition.

**Table 2:**
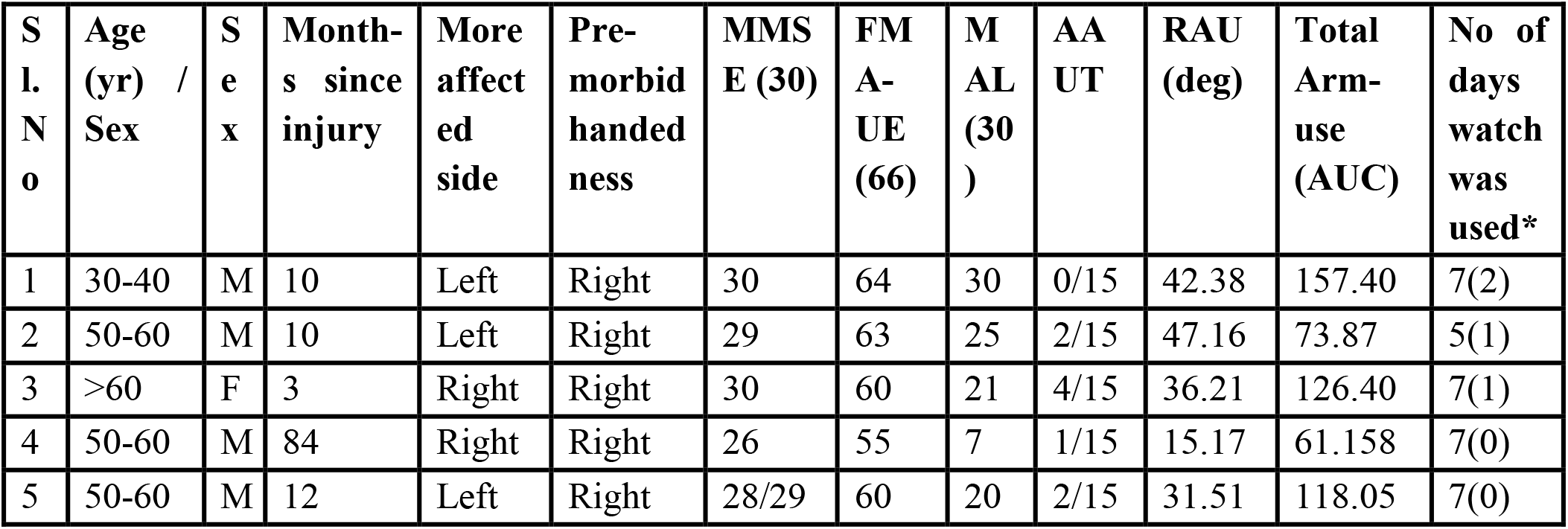
Demographic details of hemiparetic patients in the in-home study. MMSE: Mini-mental State Examination, FMA-UE: Fugl-Meyer Assessment – Upper Extremity, MAL: Motor Activity Log, RAU: Relative Arm Use, AUC: Area Under Curve, *No of additional incomplete days of recording

All patients, except P2, completed 7 days of data recording at home. On average, subjects wore the watch for about 11.91±3.96 hours each day. If there was a technical issue with the watches (e.g. improper time synchronization between the watches due to loss of power), patients informed the experimenter and the watches were replaced on the same day. In such cases, the days with incomplete recordings were not considered for analysis, and an additional day of recording was performed to complete 7 days of recording, when possible.

Fig. 3 shows the time plot of the mean arm-use of the left and right arms which are shown in red and blue, respectively, for a single day for two subjects. Patient P1 (Fig. 3(a)) who showed high overall arm-use for the two arms had a high FMA, high MAL, and low AAUT scores. For P1, despite the non-dominant side being more affected, his overall arm-use was comparable to the dominant arm. In contrast, for patient P4, the overall arm-use of the more-affected dominant right arm was much lower than the other arm; P4 had a lower FMA and MAL, and a higher AAUT. We also observed an overall reduction in the mean arm-use graphs in P4 compared to P1.

**Fig 3:**
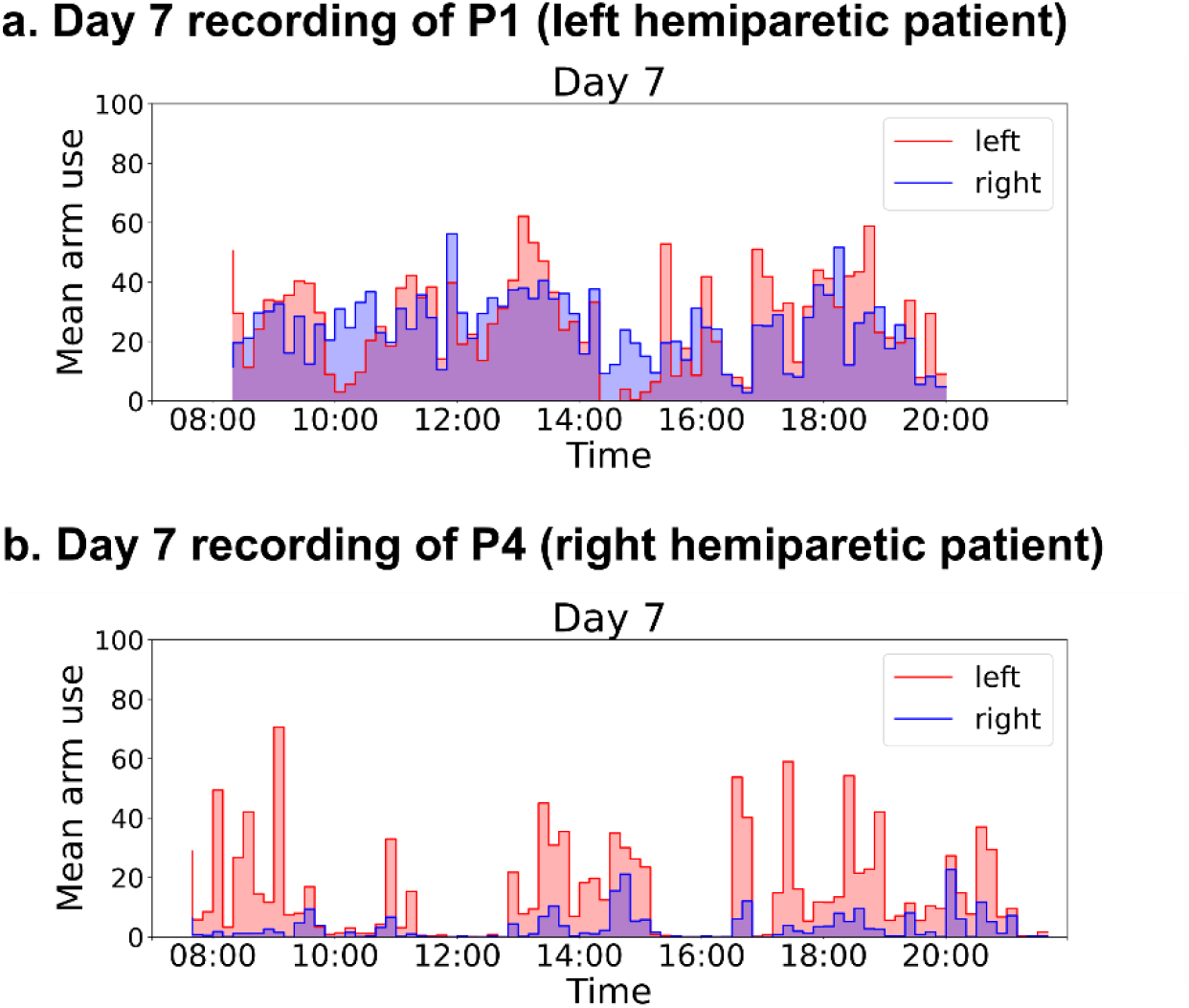
Change in arm use during a day. The red and blue colors represent left and right arm-use, respectively (a) Data from P1 (left impaired) (b) Data from P4 (right impaired). A near-normal overall activity with balanced use of both arms is seen in P1. The total activity level is less for P4 with relatively more use of the less-affected side.

Fig. 4(a)-(f) depict the arm-use scatterplots for healthy controls and patients. Fig. 4(a) shows the pooled scatterplot of arm-use data for entire recording duration from all the five healthy controls. Fig 4.(b)-(f) show the scatterplot for the five patients individually, where each plot displays the entire data collected from a patient during the 5–7 day in-home assessment. The background (in blue) in these plots, is the kernel density estimate of the controls’ scatterplot in Fig. 4(a), which serves as an indicator of expected normative behavior. This allows one to quickly identify deviations from normal arm-use behavior in a patient’s data. An asymmetry/bias in arm-use results in points clustering towards the x-axis (less affected side), and a reduction in overall arm-use leads to higher density clusters closer to the origin.

Fig. 4(a)-(f) also display the corresponding *ρ*(*ϕ*) curves plotted as a function of *ϕ*. The red curves in Fig. 4(b)-(f) are the *ρ*(*ϕ*) corresponding to individual patients, while the green curve in the background is *ρ*(*ϕ*) for all controls (same as the one in Fig. 4(a)). Two trends can be observed in the patient data compared to healthy controls:

i. **Patients have reduced total arm-use:** The *total arm-use* (area under *ρ*(*ϕ*)) for patients is lower than that of healthy controls which indicates that patients use their arms less than age-matched controls (right box-plot of Fig. 4(g)).
ii. **Some patients have asymmetric arm use**. The *ρ*(*ϕ*) curves of patients P3, P4, and P5 are skewed to the right, indicating a bias towards the use of the less-affected arm. The *relative arm-use* for these three patients is lower than that of healthy controls (bottom three points of the left box-plot of Fig. 4(g)).

**Fig 4:**
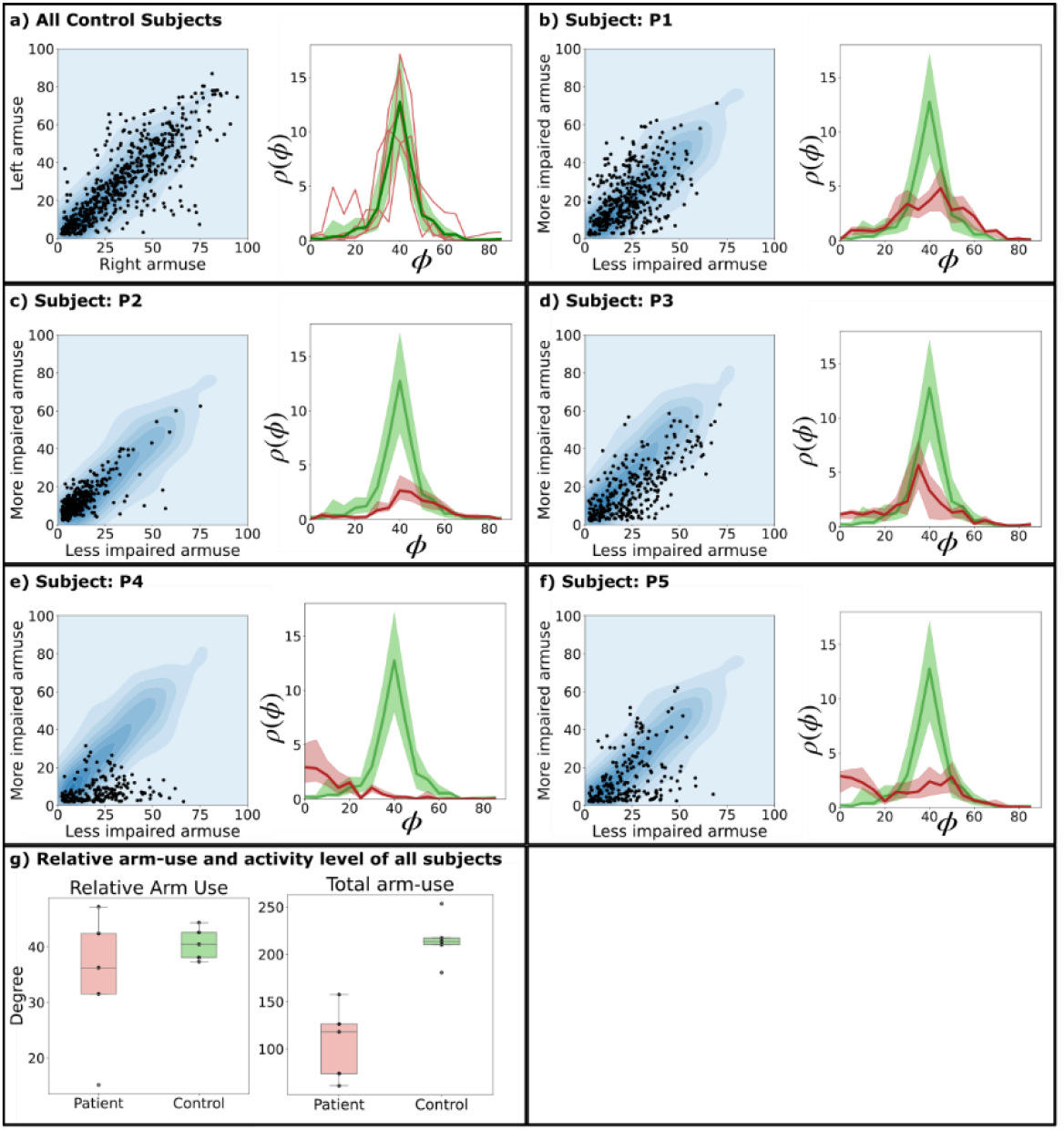
The graph shows the distribution of arm-use between the left and right arm and the *ρ*(*ϕ*) for (a) control and (b-f) patients. The control data is shown as background for reference in the scatterplots. Note that the kernel density background is mirrored about the x = y line for patients with left-hemiparesis. (g) Box plot of relative arm use and total arm-use of patients and controls

## Discussion

The current work presented: (a) the use of wearable IMUs to track relative arm-use in hemiparetic patients at home, and (b) a validation study to understand the nature of the information gathered by the gross movement score. Unlike previous work that had used acceleration thresholding^17^ or activity counting^20^, the current work used the gross movement score (*γ*) algorithm to track functional arm-use. The validation study evaluated the ability of *γ* to detect functionally relevant upper-limb movements. The in-home study with five hemiparetic patients tracked their arm-use for 5–7 days, and was used to develop appropriate analysis and visualization methods for assessing the relative arm-use. This study also helped to identify issues with using wearable technology for tracking arm-use at home.

### What does the gross movement score measure?

The *γ* forms the basis of the current work on assessing relative arm-use. Activity counting, employed in previous studies^20,30^, is highly sensitive to a wide range of movements, but is agnostic to a movement’s functional utility. On the other hand, the *γ* is a more systematic approach to detecting functional arm-use, as its algorithm exploits a common structure in functional tasks – most such tasks are performed at the level of the torso, which can be detected through orientation estimation of a wrist-worn IMU.

The *γ* has moderate agreement with functional movements and is 50–60% accurate in detecting functional movements in hemiparetic patients (Table 2). It was found to miss more functional movements of interest (false-negative rate: 34–41%) rather than detect non-functional ones (false-positive rate:6–9%). Some of the reasons for these inaccuracies include:

i. The *γ* only detects movements in a pre-defined functional range (±30° of elevation). Thus, arm movements in and out of this range during a functional task will result in fragmented *γ* which increases the false-negative rate, e.g., drinking from a cup (Fig. 2(b)), grooming, etc.
ii. It uses a velocity threshold of 15º/s to detect movements, so very slow arm movements or postures are not detected, e.g., arm postures used for object stabilization (Fig. 2(d)) or body support, writing, etc.
iii. Some of the erroneous detections by *γ* could be due to the involuntary or passive movements which are not of functional significance to the therapist.

Overall, the *γ* detects the functionally relevant movements investigated in this study with moderate accuracy, and is relatively robust to non-functional movements.

### Tracking arm-use at home

The current study identified several technical and usability issues associated using IMU-watches at homes. The following are some of the concerns raised by the study participants:

i. **Cosmetics of wearing two watch-like devices** on their wrists was one of the concerns raised by subjects. A design where one of the devices looks like a watch, while the other looks like a band might help address this issue. This feature will also reduce the chances of patients accidentally swapping the watches corresponding to the two arms.
ii. **Loss of time synchronization** between the watches because of loss of power. Sometimes when subjects forgot to recharge the watches on time, the watches would reset and lose the time synchronization. In such circumstances, the experimenters had to visit the subject at home to reset and synchronize the real-time clocks. One subject used the watches only for 5 days because he was unhappy with this technical glitch with the IMU-watches.
iii. **Lack of immediate feedback of arm-use**. The results of arm-use assessment with the watches were given to patients after analyzing the data at the end of the seven-day recording. Unanimous feedback from patients was that they would have preferred to receive daily feedback about the use of their more-impaired arm.
iv. **Lack of waterproofing** the watch casing meant patients sometimes had to remove the watch when coming in contact with water.

### Measuring, visualizing, and interpreting arm-use measures

The arm-use scatterplot approach for visualizing relative arm-use of the two upper-limbs has a similar flavour to the one proposed by Bailey et al.^20^. The scatterplot provides a measure of the overall relative arm-use in a given observation period and is devoid of any temporal information. The space occupied by the arm-use scatterplot is a square with sides of length 100 units. All straight lines of the form *y* = *mx* in this space correspond to points with a fixed ratio (*m*) between the mean arm-use of the upper-limbs on the y-axis (*𝒜_y_*) with respect to the one on the x-axis (*𝒜_x_*), i.e. 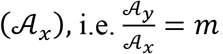. The line with *m* = 1 corresponds to points where there is equal use of both upper-limbs, while both smaller and larger slopes correspond to biased use of one of the upper-limbs. The two special cases, *m* = 0 and *m* = ∞ correspond to unilateral arm-use, i.e., only one of the upper-limbs is used during a 10min window. Care must be taken when trying to infer bilateral arm-use from this plot. We define bilateral arm-use as the simultaneous, coordinated use of the two upper-limbs to accomplish either a common goal or independent goals^31^. Thus, bilateral arm-use requires both arms’ gross movement scores to be 1, simultaneously. For example, the point **A_x_**[*k*] = 20 and **A_y_**[*k*] = 20 implies that the two upper-limbs were used for 2 min during a particular 10min window, but it does not necessarily mean they were used together to perform a bilateral task; the nature of use (uni-vs. bilateral) cannot always be exactly ascertained from mean arm-use data. This issue is similar to that of the *bilateral magnitude* proposed by Bailey et al^20^, where bilateral magnitude is computed as the sum of the activity counts of the two arms. The individual contributions of the arms cannot be determined from this sum, which was the reason for computing the *magnitude ratio* in their work^18^. The arm-use scatterplot proposed in this study is easier to interpret and quantify (e.g., through its corresponding *ρ*(*ϕ*) curve) compared to the *bilateral-magnitude* versus *magnitude-ratio* plot.

The scatterplots and their accompanying *ρ*(*ϕ*) plots allow a qualitative comparison of the arm-use behavior of patients with controls. Healthy controls used both upper-limbs almost an equal amount of time, as seen from the scatter of points about the *y* = *x* line. The RAU was 40.52º, which indicates a slight bias towards the dominant (right) side. Similar observations were made by Bailey et al.^20^. The preference for the non-dominant arm for stabilization tasks^32^ could be one of the reasons for this bias.

Patients had lower *total arm-use* than healthy controls, similar to the observations made in the Bailey et al. study^20^. The Patients P3, P4 and P5 showed a bias towards the less-affected arm as seen in the scatterplot and their corresponding *ρ*(*ϕ*) curves (Fig. 4(d)-(f)); these three patients had the lowest scores on the MAL. Patients P4 and P5 had low AAUT scores of 1 and 2 respectively, which indicate good spontaneous use of the paretic upper-limb. However, this was not reflected in the strong bias towards the less-affected side seen in their MAL or RAU scores (Table 2). This discrepancy is most likely because the hospital environment might encourage patients to choose their paretic side in the free choice condition, leading to overestimation of arm-use^3^. Arm-use of the high functioning patient P1 was very similar to that of healthy controls in terms of arm-use symmetry, but the *total arm-use* was still smaller than healthy. These trends in arm-use symmetry also have similarities to that of Bailey et al.^20^, where arm-use symmetry was slightly correlated with the ARAT score.

In addition to measures and visualization of overall arm-use and its symmetry, we strongly believe that temporal plots of arm-use, as shown in Fig. 2, are essential for understanding how the patients incorporate their upper-limbs in daily life. The temporal visualization of the mean arm- use for the two upper-limbs in Fig. 2 display variations in arm-use over time. Such plots can help identify periods of high and low upper-limb usage can give us clues about the nature of arm-use, e.g., the arm-use pattern for eating during usual meal times. Most previous studies have removed temporal dependence when obtaining an overall measure of arm-use^30^, and while visualizing arm-use data^20,30^

## Limitations

The main limitation of the current study is the small sample size of patients used for tracking arm behaviour during daily life, which does not allow us to draw general conclusions from the results. However, the current study did serve its purpose as a stepping stone towards a larger study for assessing arm-use. The measures and visualization methods proposed in the study are preliminary ideas, and the current work did not carry out a direct comparison with existing methods^17,18,30^. The *γ* estimated from IMU data is the basis of arm-use analysis in the current study. Although, a simple starting point, an analysis based only on *γ* does not make full use of the raw IMU data, which most likely contains a good amount of clinically relevant and useful information about arm-use. For example, the *γ* is a time-based measure that does not take into account the intensity of a subject’s movements (e.g., the activity counts^20^), or the temporal pattern of accelerations and angular rotation associated with different tasks. Future work must investigate the use of advanced methods for evaluating the patterns of arm-use, and its movement quality. Finally, we used the parameters proposed by Leuenberger et al.^25^ for estimating *γ* from IMU data. It is possible that patient-specific parameters could lead to better accuracy in detecting functional movements. The false negative rate of the gross movement score was about 30–40%, which is quite high. This can be addressed through appropriate changes to the hardware of the IMU-watch and to the gross movement score algorithm.

## Conclusion

The current work proposed an approach for relative arm-use assessment based on detecting functional movements, which is different from the standard activity counting approach. The work explored the feasibility of using the gross movement score extracted from wrist-worn IMUs to measure relative arm-use of community dwelling hemiparetic patients. The validation of the gross movement score found that it detects functional movements with 50–60% accuracy, and is robust to non-functional movements. The work also presented new measures and visualization methods to analyze arm-use data obtained from IMUs, which is easier to interpret. The ability to distinguish between functional versus non-functional movements allows one to analyze these components individually, and gain a deeper understanding of a patient’s arm-use pattern.

## Data Availability

Data are available on request to authors.

## Acknowledgements

We thank Dr. A.T. Prabhakar for critically discussing our work. We also acknowledge Jayashree P, Mahesh K, Jeena M and Tanya S for assistance in data collection.

## Declaration of Conflicting Interests

Dr Varadhan SKM has a financial interest in Kriya Neurotechnologies Private Limited, a company that makes IMU based Neurorehabilitation devices. The study was not funded by the company. The company had no role in the study design, data collection or analysis. All other authors declare that there is no conflict of interest.

## Funding

The authors disclosed receipt of the following financial support for the research of this article: This work was supported by the Fluid Research Grant from CMC Vellore. (grant number: IRB Min No 11303)

## Notes

### Clinical Trial

CTRI/2018/09/015648

### Author Declarations

The institutional review board of Christian Medical College Vellore, India approved this study.

